# Adaptive health technology assessments to inform oncology based priority setting in India

**DOI:** 10.1101/2023.08.01.23293485

**Authors:** Srobana Ghosh, C S Pramesh, Manju Sengar, Priya Ranganathan, Cassandra Nemzoff, Francis Ruiz, Tabassum Wadasadawala, Prakash Nayak, Jayashree Thorat, Apurva Ashok, Malkeet Singh, Abha Mehndiratta, Hiral Anil Shah

## Abstract

**Background:** Health technology assessment (HTA) is a valuable tool for informing the most efficient allocation of resources, yet is highly resource intensive limiting its applicability in practice. Adapting the HTA process by leveraging available international evidence offers a pragmatic solution to such challenges, saving time whilst still generating useful insight for decision making.

**Objective:** To develop an adaptive HTA (aHTA) framework that could be used to assess the likely cost-effectiveness of cancer treatments to inform the available entitlements under the Indian national health insurance scheme.

**Methods:** The International Decision Support Initiative worked with the National Cancer Grid in India to develop an aHTA framework which included topic selection, scoping, evidence review and appraisal to estimate the likely cost-effectiveness of cancer interventions. The evidence included international data on cost effectiveness (HTA reports and economic evaluations), price benchmarking and treatment cost estimates. Ten interventions were assessed with the newly developed framework by a working group of clinicians and health economists from both institutions.

**Results:** Of 10 interventions assessed, 9 had sufficient evidence to decide cost-effectiveness; 3 were cost-effective (1 only after a discount, 1 using the generic price), 5 were not and 1 was not cost-effective for all but was in a subgroup. A full HTA was recommended for one intervention due to uncertainty. Information on the likely cost-effectiveness, clinical benefits and treatment costs was consistently available through publicly available evidence. India on average paid almost 4 times the list price of other countries.

**Conclusion:** aHTA provides an alternative to using no economic evidence at all if a full HTA cannot be conducted. It is well-suited to cancer drugs for which there is ample available international information on cost-effectiveness. Our framework quickly generated consistent, transparent evidence to inform guidelines. The approach may be replicable in other settings in supplement to full HTA.

**Key messages:** *What is already known on this topic:* There is an enormous need for more evidence informed priority setting, however there are often significant challenges to doing so such as insufficient data, capacity and resources available. Some countries are exploring the use of rapid or adaptive methods of health technology assessment (HTA) but there is no clear guidance on the methodology that should be used.

*What this study adds:* Experience of the results of an adaptive HTA framework in practice, based on 10 oncology problems.

*How this study might affect research, practice or policy:* Other countries or institutions with limited capacity for HTA could potentially use the framework to perform their own adaptive HTA assessments.

## Introduction

India, like many other low- and middle-income countries (LMICs), has been facing an increasing double burden of disease. Whilst still experiencing high rates of communicable disease, the rates of non-communicable disease (NCDs) are also rising due to the aging population alongside the growing prevalence of risk factors such as tobacco, alcohol and obesity. Whilst cases of all NCDs are rising, the increase in cancer rates is of particular concern. The Global Cancer Observatory estimated cancer incidence in India as 1.3 million new cases in 2020, with 2.7 million 5-year prevalence cases(1).

Cancer care is costly, yet there is limited public funding for treatment (<2% of GDP)(2–4) and accessible insurance options are scarce. Estimates calculate that 25% of India’s population have had any sort of health insurance coverage(2) but the true percentage could be less. Limited public health spending by the government forces the majority of patients to seek treatment in the private healthcare sector, leading to catastrophic out-of-pocket expenditure(5, 6) and immense financial burden.

The National Cancer Grid (NCG) was established in 2012, and is the leading body for oncology treatment in India with a mandate to establish uniform standards for the prevention, diagnosis, and treatment of cancer(7) (8). It consists of a network of 287 healthcare institutions which provide up to 60% of all cancer care in India(8). The NCG also provides specialised training and education in oncology; and facilitates collaborative, translational and clinical research in cancer(9).

In 2018 the National Health Authority (NHA) of India launched the world’s largest publicly funded health insurance scheme Ayushman Bharat Pradhan Mantri Jan Arogya Yojana (AB-PMJAY) as a step towards providing universal health coverage(10, 11), offering a potential solution to escalating cancer care bills. To further improve cancer care across the country, the NHA and the National Cancer Grid (NCG) of India signed a memorandum of understanding in 2019 to develop uniform resource stratified standard treatment guidelines (STGs) for cancer services(12). These guidelines were linked to the reimbursement of the oncology treatments covered under the AB-PMJAY health benefits package (HBP).

The choice of entitlements made available under the AB-PMJAY HBP highly influences nationwide patient care and the allocation of available resources. It is therefore imperative to decide entitlements and develop the NCG guidelines based on best available evidence on safety, efficacy and cost-effectiveness. A three tier categorisation system of ‘essential’, ‘optimal’ and ‘optional’ has been developed to help categorise potential treatments(12) as, given the limits on funds, it is important that only the most cost-effective treatments be made available under AB-PMJAY, assessed through objective methods.

Health technology assessment (HTA) provides a framework to assess the value of treatments and help guide the health benefit packages under AB-PMJAY. HTA is a multidisciplinary process that uses systematic and explicit methods, including cost-effectiveness analysis amongst others, to determine the value of a health technology(13). Currently, there is a national level health technology assessment body present in India (Health Technology Assessment Programme in India (HTAIn)(14) and a large body of cost-effectiveness studies are being conducted(15), however, it is impossible to conduct a full HTA for the timely evaluation of all the potential interventions that could feed into guidelines.

Alternative methods or approaches such as adaptive HTA (aHTA) can provide the necessary evidence on the potential cost-effectiveness of selected cancer treatments in a relatively shorter time by using fewer resources and leveraging published international evidence and HTA-informed coverage recommendations(16). Adaptive HTA is a compliment to full HTA, pragmatically reducing the resource intensity of full HTA whilst producing credible findings to support decision making. By strategically adapting information from other settings, aHTA makes use of the considerable volume of available international data that already comments on the likely cost-effectiveness of treatments, in order to inform policy decisions expeditiously(16).

In order to fortify decision making for the NCG and increase the use of economic evidence, we developed a bespoke aHTA approach(17) which included rapid and targeted literature review, price benchmarking analysis and annualised treatment cost estimates, and applied it to ten oncology decision problems. The approach was designed to determine the information currently available on cost-effectiveness from international HTA agencies and the available literature, raise and question any issues of transferability of this evidence to India and to calculate the anticipated costs of treatment to develop a sense of the cost impact. Here we discuss the information gained from such an analysis and the potential policy impacts of developing an aHTA approach.

## Methods

### Overview of the aHTA framework

A technical aHTA working group consisting of health economists from the International Decision Support Initiative (iDSI)(18) and oncologists from the NCG who are familiar with full HTA processes and methods was constituted to develop and test an aHTA framework for the NCG (Figure 1). This framework includes the key processes of a full HTA: topic selection, producing a scope, evidence review, and generating a recommendation which indicates whether an intervention is potentially cost-effective, potentially not cost-effective, or if further research is needed. This recommendation is then considered in conjunction with other criteria by guideline developers as to whether to include the intervention in the STGs(12, 17). As the research was desk based, patient and public involvement was not included.

**Figure 1:**
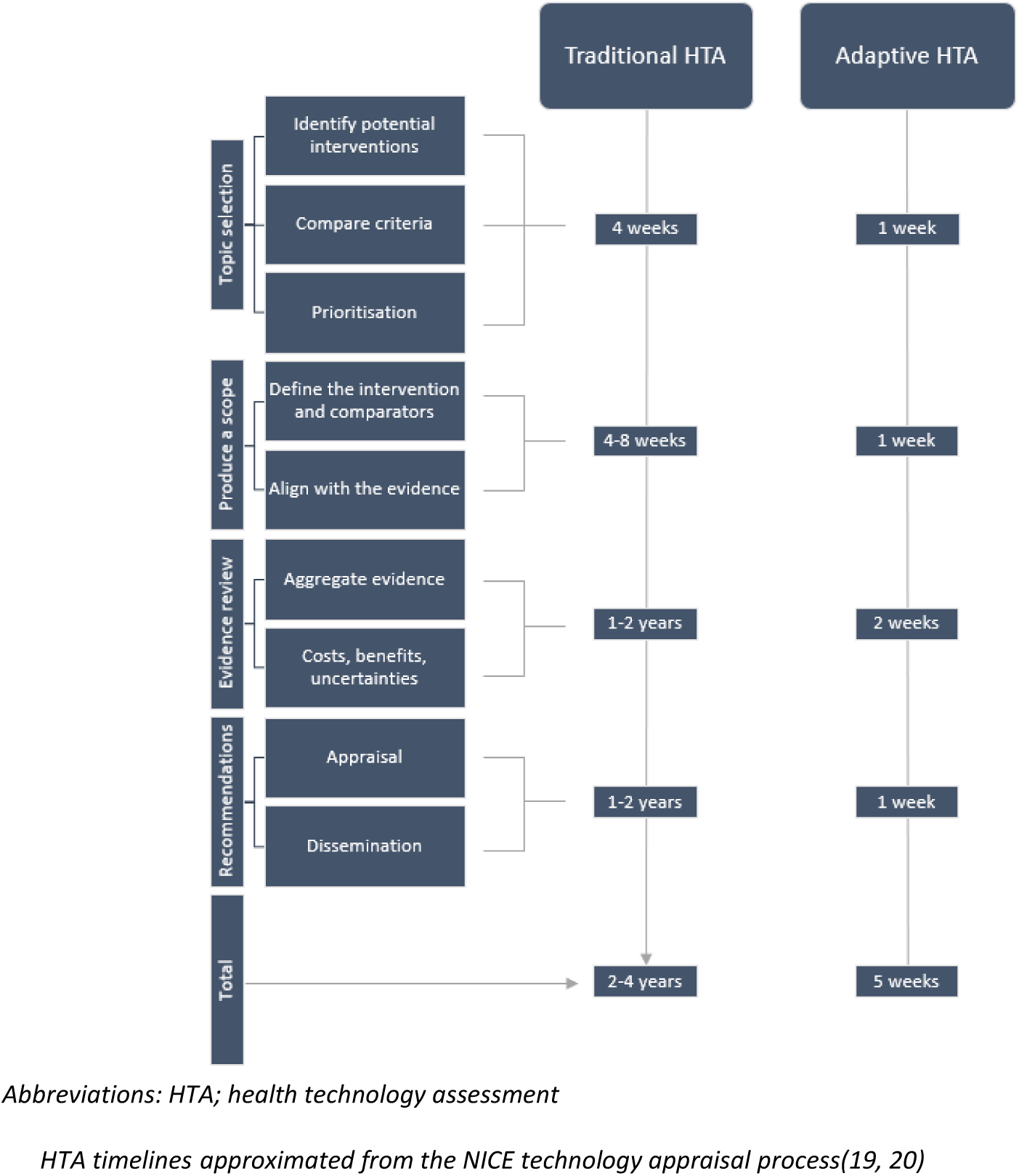
Adapted HTA process designed for the NCG.

### Topic Selection & Prioritisation

A topic selection and prioritisation process was implemented to identify, prioritise and select potential interventions for aHTA review. A list of potential interventions was identified from requests to the NCG to consider potential technologies. These interventions were then prioritised against each other based on a set of criteria, including clinical impact, treatment landscape, disease severity, disease prevalence, indicative budget impact, and availability of international data. Topics were then prioritised through a discussion process based on the evidence collated. The final list of assessments can be found in Table *1*. The process echoed that of the full HTA, but was conducted more rapidly and with a more pragmatic approach to evidence generation.

**Table 1:**
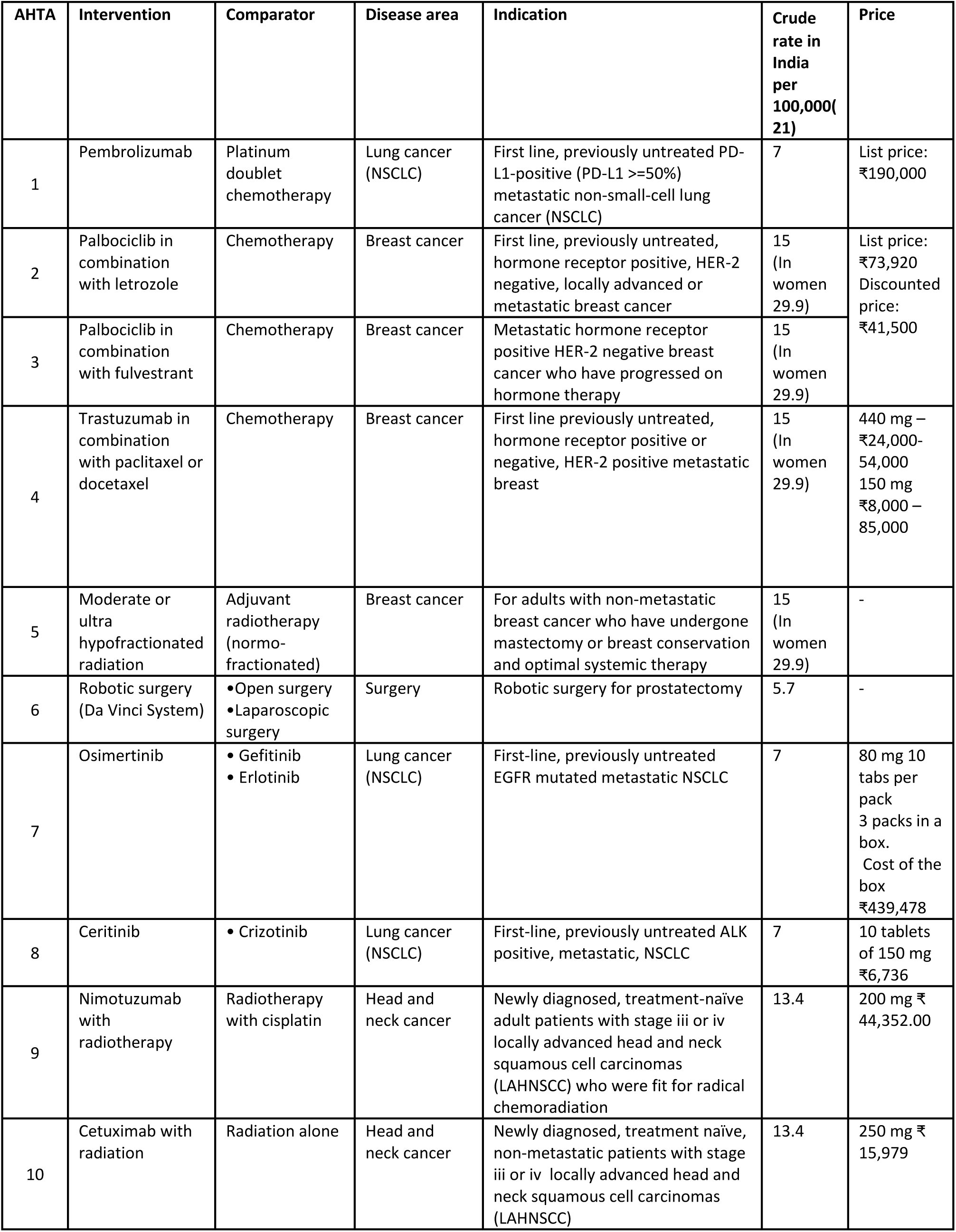
List of assessments.

### Development of a scope

The PICO (Population, Intervention, Comparator, Outcome) framework (22) was used to develop a scope for each analysis, creating a specific research question reflective of the local context derived from clinical opinion detailing the selected cancer technology in comparison to the current standard of care in India. The intervention and comparator were defined by a stated dose, with a marketing authorisation in the indication of interest. In addition to reflecting the local context, the scope was also designed to ensure that international evidence could be applied to such decision problems. As aHTA relies on leveraging international evidence, if such evidence were not available, it was decided that the topic should be reviewed through full HTA.

### Evidence review

Once a scope was produced, a rapid review of the available international evidence was performed by completing a data extraction (Figure 1) on the background, safety, clinical benefits and cost-effectiveness to ascertain how the technology was perceived in regard to its potential cost-effectiveness.

**Figure 1:**
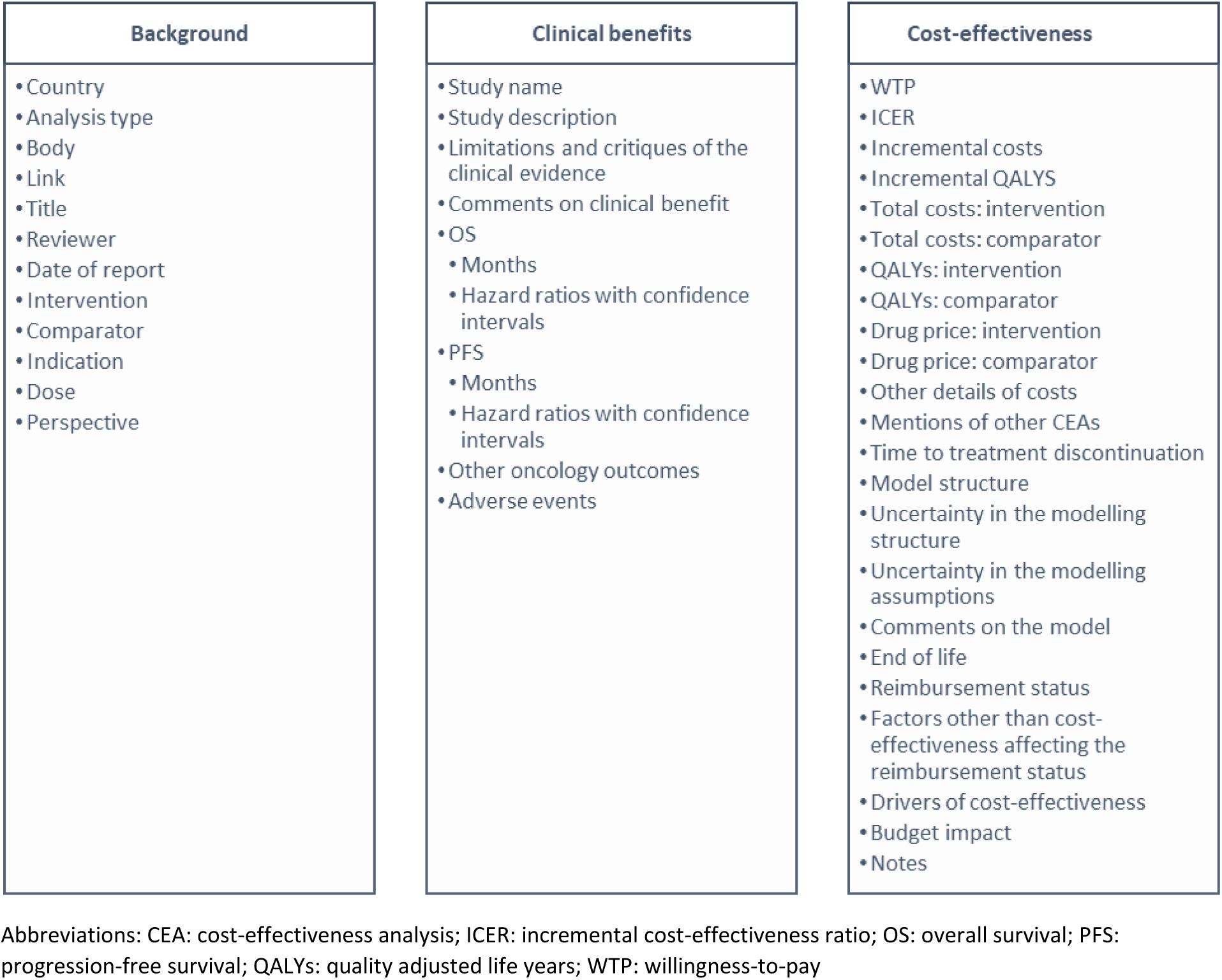
Data extraction template.

Evidence of safety and clinical benefit were extracted from the clinical trial underlying the model estimations used in international HTA agency reports or economic evaluations. Where this information was not available, as was usually the case with respect to non-pharmaceutical interventions (NPIs), evidence of clinical benefit was pragmatically sourced from peer reviewed systematic reviews and meta-analyses, clinical trials, observational studies and national clinical guidelines. Where possible, evidence that had been conducted in an Indian setting was included. Any limitations with the studies that might entail that the outputs are not generalizable to India were identified and commented upon.

Cost-effectiveness evidence was extracted from all international HTA agency reports where a decision was published or economic evaluations from a range of income settings to determine the consistency of findings, how they may change across settings, what the drivers of cost-effectiveness were and to ascertain whether the cost-effectiveness was broadly clear or at the margin.

### Price Benchmarking Analysis

To supplement the evidence review, a comparative price benchmarking analysis was conducted. The objective of the exercise was to provide a crude estimate of likely cost-effectiveness in India at the price India is paying. This was done by determining whether the intervention at the equivalent price paid in India would have been considered to be cost-effective by the other countries once adjusted for currency conversion and gross domestic product (GDP) adjusted for purchasing power parity (PPP) based on a previously published methodology(23).

**Figure 2:**
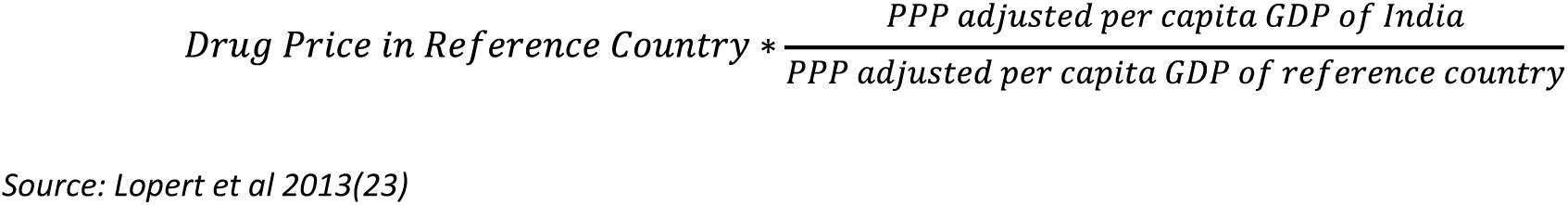
Price benchmarking formula.

The price paid in India was converted into the benchmarked country currency. This was then divided by the price the benchmarked country pays once multiplied by an adjustment factor. The adjustment factor is calculated by dividing the GDP PPP of India with that of the benchmarked country.

The approach assumed that the cost-effectiveness of a treatment could not be assured at a price higher than that paid by the reference country; therefore if India was paying a higher price, it was deemed to be not cost-effective.

### Annual Drug Cost Calculation

Whilst there was insufficient data to conduct a full budget impact analysis, given the availability of list prices for the pharmaceutical technologies, the potential annual drug cost per patient (excluding any other resource use) was estimated to gain insight into the potential cost impact of introducing the new technology under the AB-PMJAY scheme. Annual drug costs were based on the list price of the innovator and or generic, pack size, dose and number of cycles. This analysis was conducted for both the intervention and the comparator, and the difference was considered to be the treatment cost impact.

Treatment costs were also quantified as the potential fraction of the AB-PMJAY allowance that would be used up by a family if the technology were introduced, as the AB-PMJAY scheme reimbursement for any secondary or tertiary healthcare is limited to a maximum of ₹500,000 per family per year(11). If the drug has a generic version, the analysis was repeated with generic prices.

### Appraisal and recommendations

The technical aHTA working group conducted a narrative synthesis and tabulation of the results. The evidence was appraised through deliberative discussion within the group that assessed the evidence, considered transferability and raised any uncertainties. A recommendation was made by placing the intervention by means of group consensus into one of a number of pre-specified categories: “potentially cost-effective”, “potentially not cost-effective”, “potentially cost-effective for specific subgroups” or “full HTA required”. This decision could then be used by the guideline development group alongside other available evidence or considerations to judge if the treatment should be included in the STGs.

Finally, a policy brief was produced for each case study, summarising the evidence extracted from the rapid review process in addition to any price benchmarking analysis and/or annual drug cost calculations, and the intervention’s designation as to its likely cost-effectiveness.

## Results

### Summary of aHTAs

Ten aHTAs were conducted as part of the development and testing of the NCG aHTA framework. Of the ten aHTAs, the disease areas that were reviewed were breast cancer (n = 4), lung cancer (n = 3), head and neck cancer (n = 2) and prostate cancer (n = 1). Most aHTAs (n = 8) reviewed pharmaceutical technologies, but two aHTAs were specifically chosen to assess whether the aHTA framework could be applied to NPIs (e.g. devices, diagnostics, surgical procedures). Topics were all prioritised based on the criteria mentioned above. The aHTA framework evolved over time; the formalised aHTA topic selection and prioritisation process was implemented after aHTA 6, and a fuller evidence base was used from aHTA7.

### Evidence review

Evidence extraction for the first four aHTAs was limited as the full list of data extraction fields had not been devised yet; therefore the extracted data on clinical and cost-effectiveness evidence was initially sparse (Table *2*).

**Table 2:**
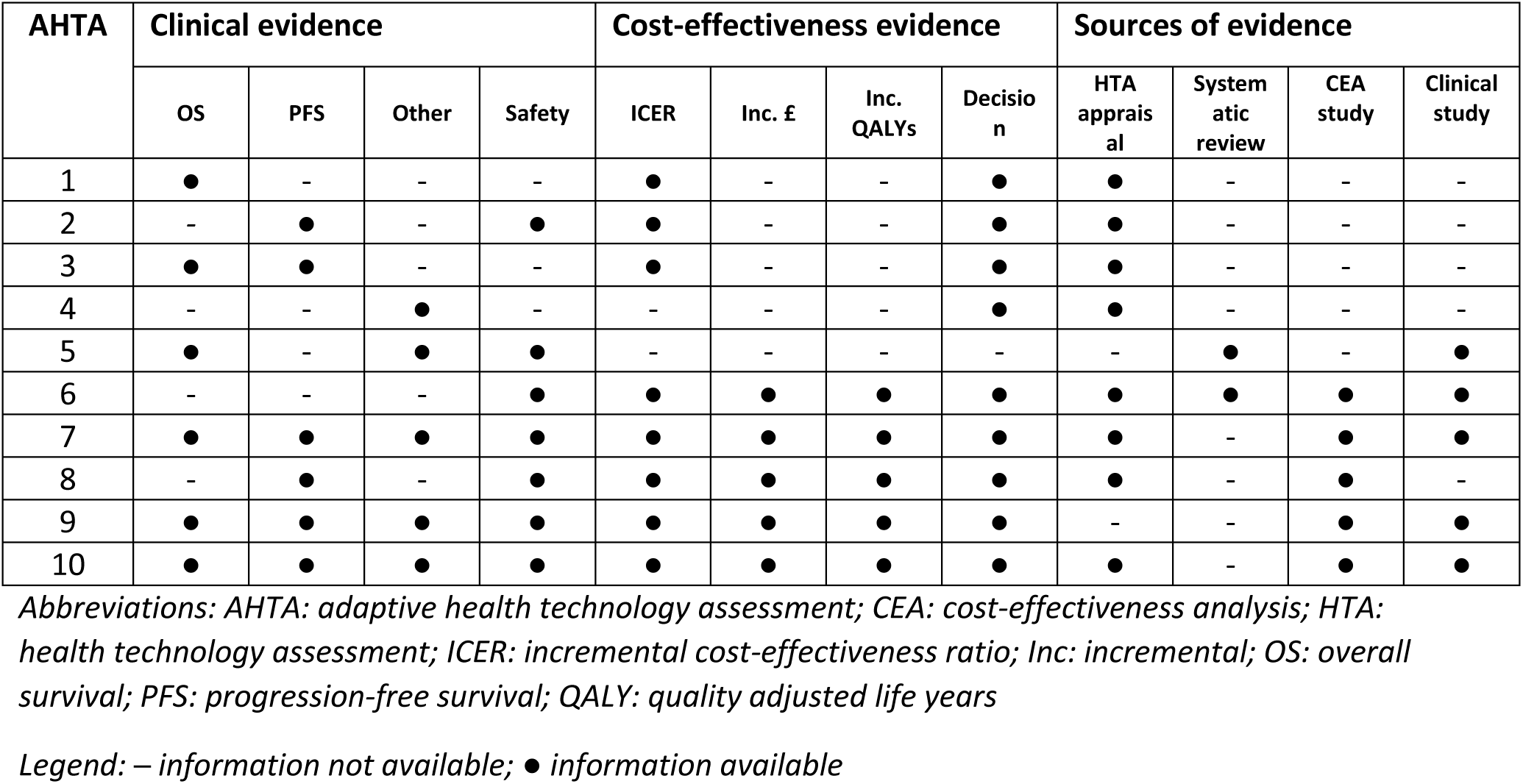
Available information.

For assessing NPIs (aHTAs 5 and 6), there was a lack of available HTA evidence therefore peer reviewed published literature was used to supplement the evidence base. Here, the framework appeared to be less suitable for assessing NPIs, but we continued including economic evaluation evidence for pharmaceutical interventions as we were able to include results from LMIC settings that may not have a formal HTA agency but were potentially more relevant for assessing cost-effectiveness in India.

As the framework developed, we updated our evidence extraction process further by increasing the number of data points that require extraction to broaden the considered evidence base included in decision making.

Of the ten aHTAs that were conducted, sufficient international evidence was extracted to assess the potential cost-effectiveness of an intervention in India for nine of them (Table 6), but aHTA 8 would benefit from a full HTA as a clear decision could not be made as to its cost-effectiveness.

### Price benchmarking

When conducting a price benchmarking analysis, we found that for the same drug at the same dosage, India commonly pays between 2-4 times more than the other countries considered in the analysis when adjusting for currency and GDP PPP(Table *3*), there was only one instance where India was paying less than another jurisdiction. The results suggest that the price India was paying was unlikely to be considered cost-effective in the benchmark countries and further discounts were needed.

**Table 3:** Price benchmarking results.

| AHTA | India price (INR) | India GDP per capita (PPP) | Benchmarked Country | Benchmarked country price | Benchmarked GDP per capita (PPP) | Benchmark country currency | PPP adjustment Factor (India GDP/ benchmark country GDP) | Currency converter rate | Indian price converted to the benchmark country price | Price paid by the benchmark country when adjusted for GDP PPP | Price Ratio |
| --- | --- | --- | --- | --- | --- | --- | --- | --- | --- | --- | --- |
| 1 | 190,000 | 7,034 | UK | 5260 | 48,710 | GBP | 0.1444 | 0.0102 | 1944 | 760 | 2.56 |
|  |  |  | New Zealand | 8000 | 43,953 | NZD | 0.1600 | 0.0203 | 3859 | 1280 | 3.01 |
|  |  |  | USA | 9724 | 65,281 | USD | 0.1078 | 0.0134 | 2545 | 1048 | 2.43 |
| 2 & 3 | 41,500 |  | UK | 2905 | 48,710 | GBP | 0.1444 | 0.0102 | 423 | 419 | 1.01 |
|  |  |  | New Zealand | 4000 | 43,953 | NZD | 0.1600 | 0.0203 | 842 | 640 | 1.32 |
|  |  |  | Australia | 4265 | 53,320 | AUD | 0.1319 | 0.0187 | 776 | 563 | 1.38 |
|  |  |  | USA | 13007 | 65,281 | USD | 0.1078 | 0.0134 | 556 | 1401 | 0.40 |
| 7 | 439,478 |  | Ireland | 6200 | 93,612 | EUR | 0.0751 | 0.0120 | 5274 | 466 | 11 |
|  |  |  | UK | 5770 | 44,916 | GBP | 0.1566 | 0.0098 | 4307 | 904 | 5 |
| 8 | 101,040 |  | UK | 4923 | 44,916 | GBP | 0.1566 | 0.0098 | 990 | 771 | 1.28 |
| 9 | 439,478 |  | Ireland | 6,200 | 93,612 | EUR | 0.0751 | 0.0120 | 5274 | 466 | 11 |
| 10 | 159,79 |  | UK | 178 | 44,916 | GBP | 0.1566 | 0.0097 | 155 | 27.89 | 6 |

N.B. Insufficient evidence was available to conduct price benchmarking on aHTAs 4-6

### Annual Drug Cost Calculation

Local drug costs could be estimated for all pharmaceutical aHTAs (Table 4), though treatment costs for NPIs could not be quantified as that would have required a bespoke costing exercise and there was insufficient data available on resource use and costs. Drug costs are taken from the time of the aHTA but some have had a discount since then. The results suggested that there was a low likelihood that any of the interventions would be considered cost-effective at the prices available. Annual drug costs across all eight pharmaceutical aHTAs comprised between 37% to 1,322% of the yearly AB-PMJAY family allowance. The majority of drugs exceeded the full AB-PMJAY family allowance. We further quantified the incremental drug cost between an intervention and its comparator where available (Table 5). All interventions had higher associated treatment costs than their comparators.

**Table 4:**
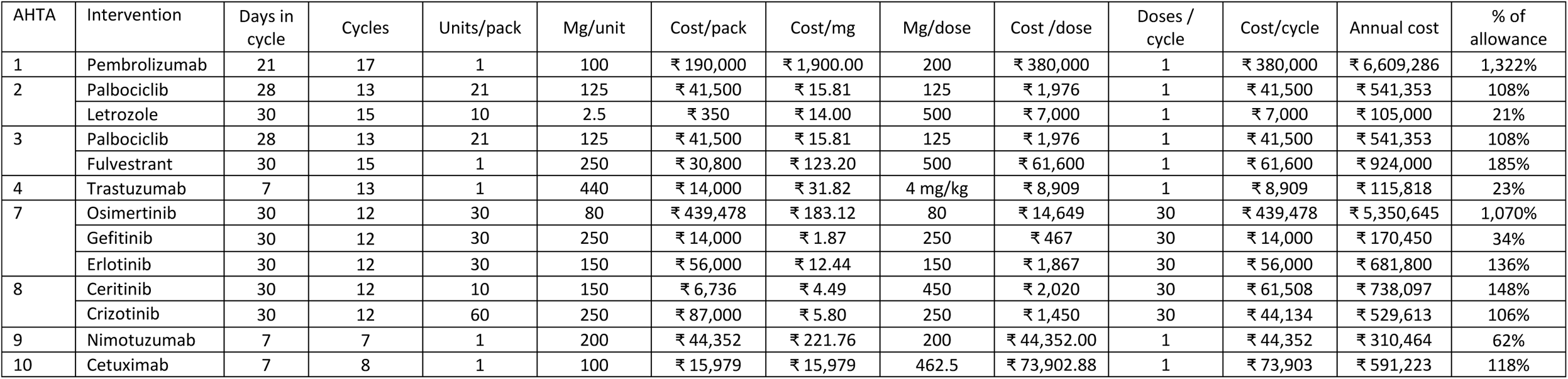
Cost of treatment.

**Table 5:** Incremental drug costs when compared to another innovator drug.

| AHTA | Intervention | Comparator | Additional treatment cost of the intervention compared to the comparator |
| --- | --- | --- | --- |
| 7 | Osimertinib | Gefitinib | + ₹ 5,180,195 |
|  |  | Erlotinib | + ₹ 4,668,845 |
| 8 | Ceritinib | Crizotinib | + ₹ 208,485 |

### Generalisability

For each AHTA, we further extracted the potential drivers of cost-effectiveness and sources of uncertainty in the health technology assessment to qualitatively assess where there might be problems of generalisability of the international HTA agency recommendation to the Indian context (Table 6). Based on our analysis and appraisal process, the aHTA working group often considered the drug price and incremental cost-effectiveness ratio to not be generalisable to the Indian context but anticipated similar levels of clinical benefit.

**Table 6:** Drivers of cost-effectiveness.

| AHTA | Likely to be cost-effective in India | Recommended in other jurisdictions |  | Drivers of cost-effectiveness |  |  |  |  |
| --- | --- | --- | --- | --- | --- | --- | --- | --- |
|  |  | Yes | No | Significant clinical benefit | Uncertain clinical benefit | Saves costs or resources | Very high costs | High willingness to pay threshold and/or commercial discount |
| 1 | No | England<br>USA | New Zealand | ✓ |  |  |  | ✓ |
| 2 | Not at the recommended price, but yes with a sufficient discount | Australia<br>England<br>New Zealand<br>Thailand<br>USA | - | ✓ |  |  |  | ✓ |
| 3 | No | England (CDF)<br>New Zealand<br>USA | Australia |  | ✓ |  |  | ✓ |
| 4 | Not at the full price but the generic price appears to be cost-effective | Australia<br>England<br>USA<br>New Zealand | - | ✓ |  |  |  | ✓ |
| 5 | Yes | No HTA Agency has appraised<br>the CEA but multiple studies found that it could be cost-effective | - |  |  | ✓ |  |  |
| 6 | No | USA | Australia-Potentially<br>Canada – Potentially<br>Canada |  | ✓ |  | ✓ |  |
| 7 | No | England<br>Ireland<br>WHO – Yes when no feasibility, cost, or affordability constraints limit the access | Canada<br>Singapore |  | ✓ |  | ✓ | ✓ |
| 8 | Full HTA needed | Canada<br>China<br>England |  |  | ✓ |  |  | ✓ |
| 9 | No | China – Yes (but not cost-effective) |  |  | ✓ |  | ✓ |  |
| 10 | Not for all patients but potentially cost-effective for a subgroup based on efficacy data | England – Yes for a subgroup of patients |  |  | ✓ |  |  |  |

Most international HTA agencies that recommended a technology used a higher willingness-to-pay threshold and had commercial arrangements in place that included significant discounts which were not applicable to India. For example in aHTA7, it was noted that all the countries that had recommended osimertinib did so at a discounted price, however the price paid in India was much higher comparatively according to the price benchmarking analysis and would comprise 1,070% of a family’s annual allowance under the AB-PMJAY. Cost offsets that were due to changes in resource use were further considered to not be replicable in India due to the differences in setting.

Importantly, the decision from the aHTA framework rarely relied on one data point, but considered the evidence base as a whole with the concerns regarding generalisability and uncertainty informing a decision on the likely cost-effectiveness offered by the intervention.

## Discussion

Sustainable universal health care requires developing frameworks for priority setting to support making difficult choices. Yet for countries such as India that have limited time, capacity and resources to undertake traditional HTA processes the current alternative is to have no economic evidence feed into decision making at all.

This is especially the case when developing standard treatment guidelines, where it would simply not be feasible to undertake “full HTAs” for all relevant intervention questions. There is therefore a strong need for a more pragmatic approach that can adapt the HTA process and leverage the existing available international evidence on cost-effectiveness to feed into decision making and help promote prioritisation in the healthcare system.

There is a precedent for using pragmatic evidence generation. Guideline developers in the past have used a range of techniques from a qualitative weighing up of costs and benefits(24) developing a best estimate of cost-effectiveness, reviewing potential differences in resource use and cost between the interventions alongside the results of the review of evidence of clinical effectiveness(25), or possibly the use of bespoke decision models to address specific questions(26). These techniques have been incorporated into the novel aHTA framework to develop a systematic process that provides consistency, transparency and rigour to the evidence used in every analysis.

### Suitability of aHTA

Using ten case studies, we showcase a novel aHTA framework developed by the NCG to consider economic evidence within oncology based STGs and AB-PMJAY HBPs. Through adapting the traditional HTA framework to fit with the NCG capabilities and needs, we found that the time taken to build capacity and conduct an aHTA was significantly shorter than the traditional HTA process (one month instead of two years), which allowed for many more interventions to be assessed in the time it takes to conduct one full HTA. If the results of the aHTA could be used for decision making, it would save considerable time and resources. The aHTA approach adopted here offers a credible approach to introduce cost-consciousness in the clinical guideline development process, especially for pharmaceutical products.

Based off these ten case studies, aHTA is suitable to inform price negotiations or rapidly identify which services offer the poorest value for money. aHTA can also be a valuable tool for assessing incremental additions to health benefits packages or revising existing packages by informing which services should be disinvested or should be targeted to certain subgroups to improve the financial sustainability of any national health insurance scheme. aHTA can provide an explicit evidence base for quickly determining when an intervention is highly likely to not be considered cost-effective. Finally, aHTA could also be incorporated into the traditional HTA process as a topic filter to help ensure that time and resources are saved for only the high priority technologies where there is significant uncertainty and the cost-effectiveness is marginal.

Finally, results suggest that aHTA is highly suited to technologies for which there is available evidence of cost-effectiveness from countries that have made reimbursement decisions informed by published evidence. This typically means pharmaceutical technologies (i.e. therapeutics). In this instance, a credibly rapid assessment can be made of the technology’s relative value for money, especially if there is broad consistency in recommendations across jurisdictions.

### Data availability and generalisability

For all the aHTAs, the clinical benefits were the most consistently available data point in the international literature(27), and were mostly likely to be considered to be generalisable to the Indian population based on the feedback from clinical experts. Involving clinicians in the aHTA was key in assessing transferability, capturing standard practice in India and identifying any limitations with the underlying studies that might reduce the generalisability of the results.

Estimates of cost-effectiveness from international studies were not considered to be directly generalisable to India due to the differences in prices, resource use, and setting. However, it was informative to understand what the drivers of cost-effectiveness were in these settings in order to see if the same or similar conditions applied in India. It was also important for the aHTA process to highlight areas of uncertainty in the evidence, including how confident HTA agencies were in their decision, if the decision was at the margin (i.e. borderline cost-effective), where there were points of doubt, and if there were any considerations outside of cost-effectiveness that influenced the final coverage decision. Involving health economists with experience of HTA helped to make this assessment but having clinicians with a deep knowledge of health economics is crucial. Investing in developing clinician capacity to understand and interpret health economic data will be vital to the success of any aHTA framework.

As there is considerable uncertainty introduced by aHTA due to the trade-offs between rigour and time, the development of this bespoke aHTA framework intentionally did not include data generation or use local data as this would require a level of scrutiny for which there was no time and capacity to address. Drug costs were the only local data used within this framework as they could be integrated and generalised nationally with a reasonably high level of confidence and provided some sense of the cost impact of introducing the technology. However, other cost offsets were not included as this would have required estimating costs and healthcare resource use where there was inadequate capacity and data to do so. Future research areas should aim to adapt the aHTA framework to include rapid and pragmatic ways to perform costing analysis using existing cost data sources in India(28).

### Global applicability of aHTA

The need for increasing the amount of economic evidence used in decision making is global and whilst there is no internationally defined aHTA framework, there are multiple countries that are fast tracking or exploring pragmatic methods of generating HTA evidence (29), suggesting its growing importance. The methodology used by different countries appears to reflect the capabilities and capacities of the local contexts, and can involve rapid reviews, transferring HTA reports to local contexts or running streamlined economic evaluations. The NCG framework includes elements of these methods but is rooted in a qualitative assessment of international recommendations which could be used by other regions in any indication. For example other governments could use aHTA to inform their HBPs, or clinicians from other contexts could use aHTA to inform clinical guidelines, or supranational organizations could use aHTA to inform their guidance documents.

### Limitations

Developing the aHTA framework posed several inherent challenges. Firstly, the aHTA approach described here cannot be implemented for treatments where international evidence is non-existent or where research questions do not align across jurisdictions, literature or available guidelines. It may have limited value for non-pharmaceutical interventions or drugs that have not been well studied. However, for most pharmaceutical drugs, particularly in oncology, there appeared to be substantial evidence across HTA agencies and published literature to provide timely insight into the research questions.

Secondly, cost offsets such as changes in resource use are often considered a key driver of cost-effectiveness in international appraisals, but were unlikely to be considered generalisable to the Indian context, therefore the impact on the potential cost-effectiveness was unknown. Such data are not included within our aHTA framework as no local data was available outside of drug costs that was both considered robust and could be accessed in a timely manner; therefore its inclusion would have added more uncertainty than could be accounted for in the present methodology. However, the framework could be expanded to include such data as and when it becomes available.

Finally, the NCG aHTA framework is a consultative review and a qualitative process that requires assessment and discussion. The process is reliant on a qualitative assessment to understand flaws in the underlying studies, inaccuracies in the international modelling or transferability issues. As the underlying evidence has been rigorously vetted by the international HTA agency, a reliance was made on their own assessment as to the robustness of the evidence. Future aHTA efforts could consider developing checklists of transferability or methods to assess transferability and generalisability of international evidence(30). However this would add additional analytical time and effort and the additional value that such process would offer should be considered.

## Conclusion

Greater cost-consciousness of interventions can create more efficiency and achieve more value in the health system. HTA evidence is still the most rigorous tool for priority setting and countries are urged to develop their HTA systems further, however if HTA evidence cannot be generated within the necessary timeframe, aHTA provides a reasonable alternative to using no economic evidence at all. It should be considered a supplementary process, not a substitute. Ultimately, there is a trade-off between the need for evidence and the available capacity and time to generate said evidence that aHTA can assist with. However, it is important to have a consistent, transparent and rigorous process that clearly defines what evidence included to determine cost-effectiveness.

The development and implementation of the aHTA framework within the NCG should be viewed as a successful initiative that can be expanded to other indications and contexts to support countries and institutions towards universal health coverage.

## Funding

This work was supported by the Bill & Melinda Gates Foundation Grant no: INV-003239.

## Competing Financial Interests

HS contributed to this study whilst being employed for the Center for Global Development. HS is now an employee for GSK and holds shares in the GSK group of companies. No other authors declare a competing interest.

## Acknowledgements

The authors thank Javier Guzman at the Center for Global Development for valuable comments.

## Contributions

Below is the detailed breakdown of their contribution.

- Concept and design: SG, FR, AM, HAS
- Acquisition of data: SG, MS, PR, CN, FR, TW, PN, JT, AA, MS, AM, CSP, HAS
- Analysis and interpretation of data: SG, MS, PR, CN, FR, TW, PN, JT, AA, MS, AM, CSP, HAS
- Drafting of the manuscript: SG, HAS
- Critical revision of paper for important intellectual content: SG, MS, PR, CN, FR, TW, PN, JT, AA, MS, AM, CSP, HAS
- Statistical analysis: SG, HAS
- Obtaining funding: FR, AM
- Supervision: FR, AM, HAS

## Data Availability

All data used within this analysis were from publicly available sources and can be provided upon suitable request.

## Ethics Approval: Human Participants

This study does not involve human participants. No ethics approval required.

